# Genome-wide association study of multiethnic non-syndromic orofacial cleft families identifies novel loci specific to family and phenotypic subtypes

**DOI:** 10.1101/2021.09.20.21263645

**Authors:** Nandita Mukhopadhyay, Eleanor Feingold, Lina Moreno-Uribe, George Wehby, Luz Consuelo Valencia-Ramirez, Claudia P. Restrepo Muñeton, Carmencita Padilla, Frederic Deleyiannis, Kaare Christensen, Fernando A. Poletta, Ieda M Orioli, Jacqueline T. Hecht, Carmen J. Buxó, Azeez Butali, Wasiu L. Adeyemo, Alexandre R. Vieira, John R. Shaffer, Jeffrey C. Murray, Seth M. Weinberg, Elizabeth J. Leslie, Mary L. Marazita

## Abstract

Orofacial clefts (OFCs) are among the most common craniofacial birth defects and constitute a high public health burden around the world. OFCs are phenotypically heterogeneous, affecting only the lip, only the palate, or involving both the lip and palate. Cleft palate alone is demonstrably a genetically distinct abnormality from OFCs that involve the lip, therefore, it is common to study cleft lip (CL) in combination with cleft lip plus cleft palate (CLP) as a phenotypic group (i.e. cleft lip with or without cleft palate, CL/P), usually considering CLP to be a clinically more severe form of CL. However, even within CL/P, important genetic differences among subtypes may be present. The Pittsburgh Orofacial Cleft (Pitt-OFC) multiethnic study is a rich resource for the study of non-syndromic OFC, comprising a large number of families (∼12,000 individuals) from multiple populations worldwide: US and Europe (whites), Central and South America (mixed Native American, European and African), Asia, and Africa. In this study we focused on the CL/P families from this resource grouped into three non-overlapping family types: those with only CL affected members, only CLP affected members, or both CL and CLP. In all, seven total subtypes besides the combined CL/P phenotype, were defined based on the cleft type(s) that were present within pedigree members. The full sample for these analyses includes 2,218 CL and CLP cases along with 4,537 unaffected relatives, as well as 2,673 pure controls with no family history of OFC. Genome-wide association analyses were conducted within each subset, as well as the combined sample. Five novel genome-wide significant associations were observed: 3q29 (rs62284390, p=2.70E-08), 5p13.2 (rs609659, p= 4.57E-08), 7q22.1 (rs6465810, p= 1.25E-08), 19p13.3 (rs628271, p=1.90E-08) and 20q13.33 (rs2427238, p=1.51E-09). In addition, five significant and four suggestive associations confirmed regions previously published as OFC risk loci - *PAX7, IRF6, FAM49A, DCAF4L2*, 8q24.21, *ARID3B, NTN1, TANC2* and the *WNT9B*:*WNT3*gene cluster. At each of these loci, we compared effect sizes of associated SNPs observed across subtypes and the full sample, and found that certain loci were associated with a specific cleft type, and/or specific family types. Our findings indicate that risk factors differ between cleft and family types, but each cleft type also exhibits a certain degree of genetic heterogeneity.

**AUTHOR SUMMARY:** Orofacial clefts are common birth defects. Clefts often run in families, but their genetic basis is still an active area of investigation. In this study, we use an innovative approach to identify shared and unique genetic risk factors between two types of orofacial clefts - cleft lip and cleft lip plus cleft palate, by taking the patterns of different cleft types reported in families into account. Our study provides new insights into previously known genetic risk factors, but also identifies novel genetic regions that differentially impact the risk of developing cleft lip versus cleft lip plus cleft palate. This study contributes to the growing evidence that different sets of genes impact different forms of clefting and highlights the importance of incorporating information about familial affection patterns into analyses.

## INTRODUCTION

Orofacial clefts (OFCs) are among the most common birth defects worldwide. The physical health effects of OFCs pose social, emotional and financial burdens on affected individuals and their families [1-3], despite therapies such as surgical treatments, ongoing orthodontia, speech therapy etc. that are available to reduce these burdens. Similar to other birth-related malformations, there are disparities in access to the complex medical and surgical therapies for OFCs[4]. A variety of studies have reported a reduced quality of life for children with OFC [5], as well as a higher risk of certain types of cancers in adulthood [6-8]. Thus, identifying etiologic factors responsible for OFCs is a very important tool for determining risk, designing prevention methods, and determining the extent of therapeutic and social support needed by individuals with OFCs and their families.

OFCs are heterogeneous with varying manifestations and severity but are typically categorized into three subtypes: cleft lip alone (CL), cleft palate alone (CP), and cleft lip plus cleft palate (CLP). These can be syndromic (i.e. part of a spectrum of multiple defects due to a single cause), but the majority, about 70% of CL with or without CP (CL/P) and 50% of CP, are non-syndromic (i.e. the only defect present without any other detectable cognitive or structural abnormality) [9]. Many of the genes responsible for Mendelian forms of syndromic OFCs have been identified (OMIM, https://www.omim.org/search/advanced/geneMap) as have some teratogenic causes (ref?). In contrast, our understanding of the genetic causes of non-syndromic OFCs (nsOFCs) remains incomplete due to the complex nature of these defects, despite studies over a number of years [10, 11]. Not only are there differences in birth prevalence around the world with respect to any nsOFC, the prevalence of the various subtypes (CL, CLP, CP) also varies substantially, suggesting etiological differences in the genetic factors giving rise to these different forms of nsOFC. These differences likely reflect the fact that human craniofacial development is a multi-stage process involving complex interactions between genetic and environmental factors [11].

Historically, CL and CLP have been treated as variants of the same defect based on embryological origins of the upper lip and secondary palate, with CLP being considered a more severe form of CL [14]. Analysis of recurrence risk among siblings have shown that the cross-subtype recurrence risk ratio between CL and CLP is higher than between CP and either CL or CLP [15], and analyzing the composite phenotype with lip involvement (CL/P) within association analyses have resulted in consistently stronger signals, than analyzing all three (CL, CLP, CP) as a combined phenotype. Therefore, CP has been treated as being genetically distinct from nsOFCs involving the lip. More recently, it has been shown that CL and CLP have shared and unique etiological factors, therefore, recent genetic studies have focused on investigating etiological differences between CL and CLP, including both candidate gene approaches [16, 17] as well as genome-wide association study (GWAS) approaches [18-20].

Our current study focuses on nsOFC and investigates whether CL is etiologically different from CLP by considering the types of clefts segregating within families. This family-type based approach was previously used for genome-wide linkage-analyses [21], but has not been employed for GWASs. Following a methodology similar to the prior family-based analysis for partitioning families [21], we created several GWAS samples and phenotypes, as defined in the Terminology section below, and described in detail in Methods. This approach stands in contrast to previous GWASs, including those utilizing Pittsburgh Orofacial Cleft Study (Pitt-OFC) participants [12, 13] that have focused only on the *individual* subjects’ cleft types (see e.g. table in [11]). The Pitt-OFC resource is a rich collection of nsOFC families across multiple racial/ethnic groups, including simplex, multiplex, and extended pedigrees (∼12,000 participants) with precise and detailed information on the types of nsOFC observed within multiple generations of the relatives of the probands. This resource is therefore well suited to investigating differences between the genetic etiology of CL vs. that of CLP. Study samples were genotyped on a custom whole genome genotyping array, followed by imputation using the 1000 Genomes Project reference panel (phase 3). In our current study, we selected families containing one or more individuals affected with CL and/or CLP, excluding families with only CP.

### Terminology

Three non-overlapping types of families were considered: **CL** – all affected members have CL; **CLP** - all affected members have CLP; and **CL+CLP** - families containing CL as well as CLP affected members. Further, **CL+** designates the union of CL and CL+CLP families, **CLP+** designates the union of CLP and CL+CLP families, and **POFC** is used to designate the union of **CL, CLP** and **CL+CLP**. Eight phenotype analysis subgroups were then defined on these family types for analysis. The following designations list the OFC phenotype analysis subgroups with a subscript for the family type(s) included in each: **CL/P**_**[POFC]**_ is the full sample analyzed by assigning a positive affection status to both CL- and CLP-affected subjects. **CL**_**[CL]**_ is the GWAS sample and phenotype including pedigrees with only CL-affected (no CLP-affected) members, and **CLP**_**[CLP]**_ only CLP-affected (no CL-affected). **CL/P**_**[CL+CLP]**_ is the sample and phenotype consisting of pedigrees with both CL and CLP affecteds, assigning a positive affection status to both CL and CLP members. Similarly, **CL**_**[CL+CLP]**_ and **CLP**_**[CL+CLP]**_ are samples also consisting of pedigrees with both CL and CLP affecteds, but with only CL members set to affected (CLP members excluded), or only CLP members set to affected (CL members excluded) respectively. Finally, **CL**_**[CL+]**_ and **CLP**_**[CLP+]**_ are samples consisting of the CL+ or CLP+ family groups; respectively, but with only CL members set to affected (CLP members excluded), or only CLP members set to affected (CL members excluded). Fig 1 shows the GWAS sample definition and phenotype assignment used in this study. Table 1 lists selected prior studies of OFC types on Pitt-OFC subjects that most closely resemble the subset and phenotypes analyzed in our study.

**Fig 1.**
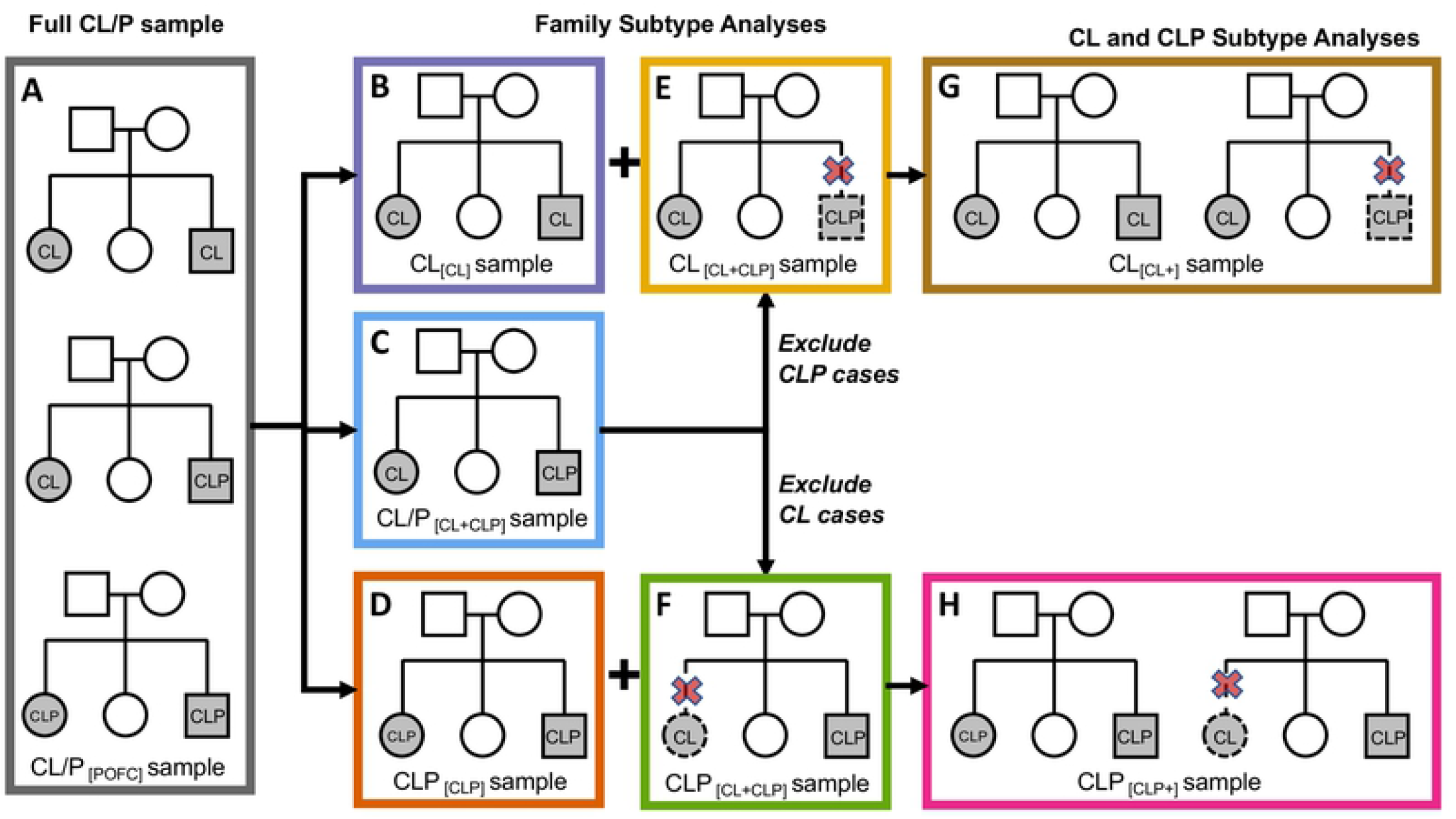
Creation of analytical subsets and phenotype assignment for GWAS. Each colored rectangle is a GWAS phenotypic subset; included pedigree type(s) shown for each subset; shaded squares and circles indicate participants with an OFC; shaded circles and squares with solid outlines indicate **affected** subjects; unshaded squares and circles with solid outlines represent **unaffected** subjects; circles and squares with dotted outlines represent pedigree members **excluded** from the GWAS; designations for OFC phenotype analysis subgroups including a subscript for the family type(s) are: (A) **CL/P**_**[POFC]**_: full set of [CL], [CLP] and [CL+CLP] pedigrees, CL and CLP members set to affected; (B) **CL**_**[CL]**_: in [CL] pedigrees, CL members are set to affected; (C) **CLP**_**[CLP]**_, in [CLP] pedigrees CLP members are set to affected; (D) **CL/P**_**[CL+CLP]**_, in [CL+CLP] pedigrees, CL and CLP members are set to affected; (E) **CL**_**[CL+CLP]**_, in [CL+CLP] pedigrees, CL members set to affected, CLP members excluded; (F) **CLP**_[CL+CLP]_, in [CL+CLP] pedigrees, CLP members are set to affected and CL members excluded; (G) **CL**_**[CL+]**_, in [CL+] pedigrees (i.e. [CL] plus [CL+CLP] pedigrees), CL members are set to affected and CLP members excluded; (H) **CLP**_**[CLP+]**_, in [CLP+] pedigrees (i.e. [CLP] plus [CL+CLP] pedigrees), CLP members are set to affected and CL members excluded. Note: Affected sibships are shown as examples – data includes other pedigree types including multi-generational pedigrees.

**Table 1.** Comparison of previous published analyses on Pitt-OFC

| Prior Study | Study type/goal | Approach | Correspondence to current study subsets |
| --- | --- | --- | --- |
| Marazita et al. 2009 [21] | Genome-wide linkage, fine-mapping | Parametric linkage (HLOD) and FBAT | $\mathbf{CL}/\mathbf{P}_{[POFC]}$, $\mathbf{CL}_{[CL]}$, $\mathbf{CLP}_{[CLP]}$ |
| Leslie et al. 2017 [13] | GWAS | TDT, case-control association and meta-analysis | $\mathbf{CL}/\mathbf{P}_{[POFC]}$ |
| Carlson et al. 2019 [16] | Heterogeneity within OFC in targeted gene regions | GWAS, meta-analysis, and heterogeneity Q-statistic with permutation testing for significance | $\mathbf{CL}_{[CL+]}$, $\mathbf{CLP}_{[CLP+]}$ |

Since the degree of OFC risk at certain susceptibility loci varies with ancestry [22], the effect of ancestry was incorporated into our analyses. The four ancestry groups used to classify study participants are AFR (African ancestry), ASIA (Asian ancestry), EUR (white, European ancestry) and CSA (Central and South American ancestry). EAF is used to denote the effect allele frequency within a specified subset of participants. LD r^2^ is used to denote linkage disequilibrium between variants as observed within the POFC sample.

## RESULTS

In our study, GWASs of eight separate phenotypes were run on eight corresponding phenotypic subsets created by grouping the POFC pedigrees based on the type of OFCs (CL and/or CLP) observed within those pedigrees. The full sample was analyzed for the CL/P phenotype (CL/P_[POFC]_), and seven other phenotype/family groups, CL_[CL]_, CLP_[CLP]_, CL/P_[CL+CLP]_, CL_[CL+CLP]_, CLP_[CL+CLP]_, CL_[CL+]_ and CLP_[CLP+]_ were defined, and analyzed using GWASs. For each phenotype, pedigrees were further grouped according to their population ancestry groups, and GWASs run separately within each group. Subsequently, association outcomes for the ancestry groups were meta-analyzed to determine association for each of the eight phenotypic subsets. The procedure followed for creating and analyzing the eight phenotypic subgroups is described in the Methods section. Genome-wide meta-analysis resulted in several significant and suggestive associations, both at previously reported OFC loci, and five novel regions.

### Significant and suggestive loci identified by meta-analysis

Meta-analysis over the ancestry groups for each of the eight phenotypes resulted in fourteen unique loci of interest. These included five novel loci with genome-wide Bonferroni significant meta-analysis p-values (p < 5.0e-08) and an additional nine known OFC loci with p-values below 1.0E-06. Table 2 lists the most significant meta-analysis p-value, effect size (expressed as betas), 95% CI of the effect size, and the variant positions that showed significant (p < 5.0e-08) or suggestive (p < 1.0e-05) associations. Supplementary Table S1 provides more detailed information for all variant positions corresponding to the p-values shown in Table 2, such as RS numbers, base pair positions, and effect allele frequencies (EAFs) within the affected subjects included for GWAS of that phenotype.

**Table 2.**
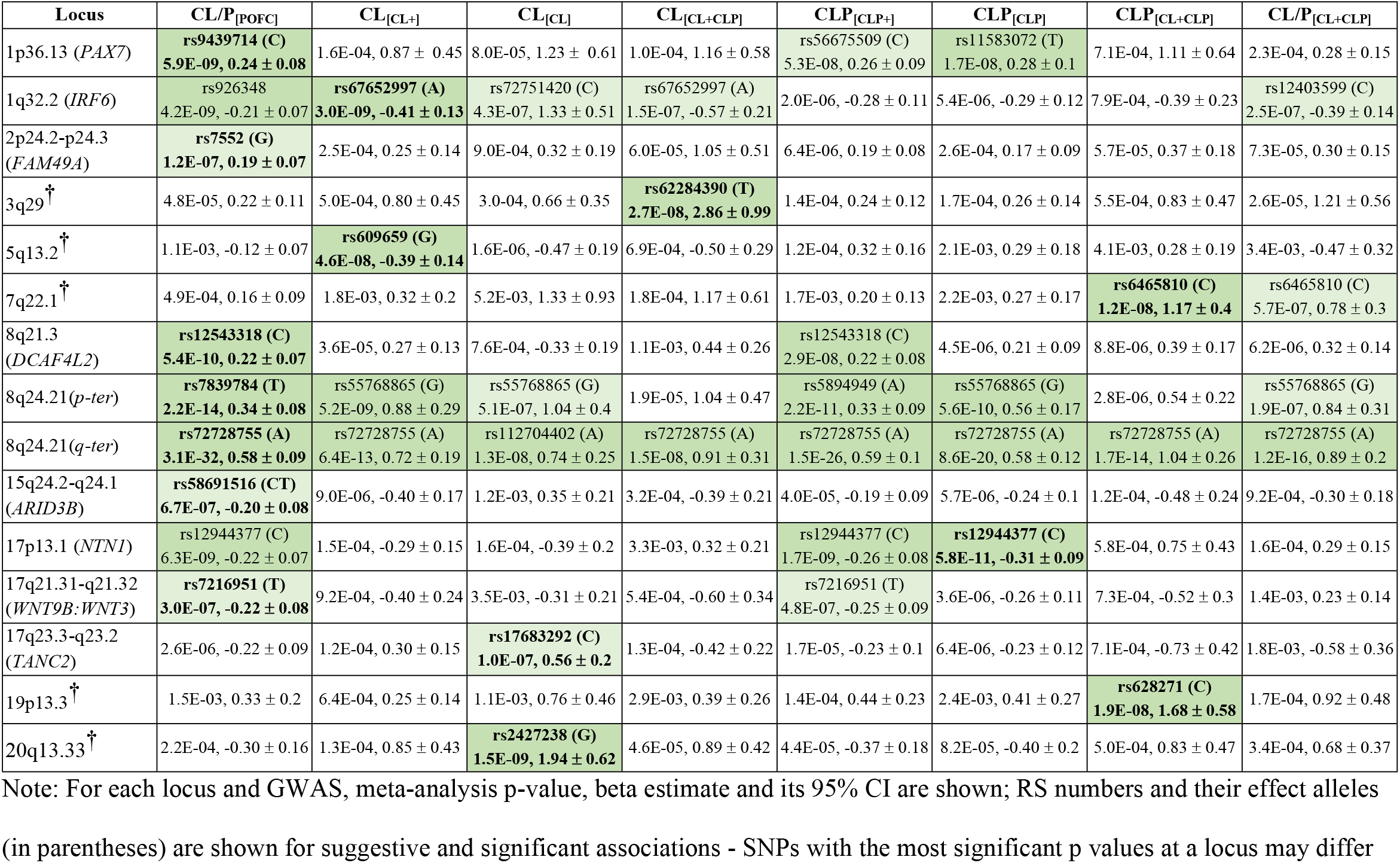
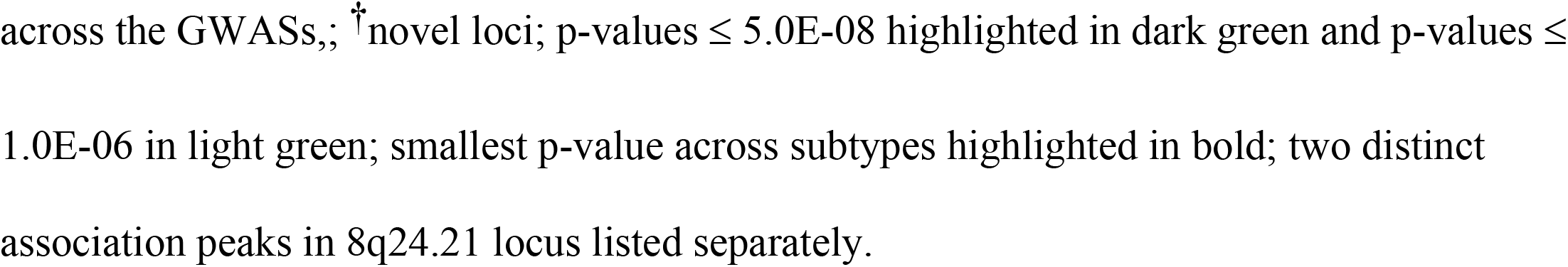
Loci with meta-analysis p-value ≤ 1.0E-06 in one or more GWASs

| Locus | CL/P<sub>[POFC]</sub> | CL<sub>[CL+]</sub> | CL<sub>[CL]</sub> | CL<sub>[CL+CLP]</sub> | CLP<sub>[CLP+]</sub> | CLP<sub>[CLP]</sub> | CLP<sub>[CL+CLP]</sub> | CL/P<sub>[CL+CLP]</sub> |
| --- | --- | --- | --- | --- | --- | --- | --- | --- |
| 1p36.13 (*PAX7*) | **rs9439714 (C)**<br>**5.9E-09, 0.24 ± 0.08** | 1.6E-04, 0.87 ± 0.45 | 8.0E-05, 1.23 ± 0.61 | 1.0E-04, 1.16 ± 0.58 | rs56675509 (C)<br>5.3E-08, 0.26 ± 0.09 | rs11583072 (T)<br>1.7E-08, 0.28 ± 0.1 | 7.1E-04, 1.11 ± 0.64 | 2.3E-04, 0.28 ± 0.15 |
| 1q32.2 (*IRF6*) | rs926348<br>4.2E-09, -0.21 ± 0.07 | **rs67652997 (A)**<br>**3.0E-09, -0.41 ± 0.13** | rs72751420 (C)<br>4.3E-07, 1.33 ± 0.51 | rs67652997 (A)<br>1.5E-07, -0.57 ± 0.21 | 2.0E-06, -0.28 ± 0.11 | 5.4E-06, -0.29 ± 0.12 | 7.9E-04, -0.39 ± 0.23 | rs12403599 (C)<br>2.5E-07, -0.39 ± 0.14 |
| 2p24.2-p24.3<br>(*FAM49A*) | **rs7552 (G)**<br>**1.2E-07, 0.19 ± 0.07** | 2.5E-04, 0.25 ± 0.14 | 9.0E-04, 0.32 ± 0.19 | 6.0E-05, 1.05 ± 0.51 | 6.4E-06, 0.19 ± 0.08 | 2.6E-04, 0.17 ± 0.09 | 5.7E-05, 0.37 ± 0.18 | 7.3E-05, 0.30 ± 0.15 |
| 3q29<sup>†</sup> | 4.8E-05, 0.22 ± 0.11 | 5.0E-04, 0.80 ± 0.45 | 3.0E-04, 0.66 ± 0.35 | **rs62284390 (T)**<br>**2.7E-08, 2.86 ± 0.99** | 1.4E-04, 0.24 ± 0.12 | 1.7E-04, 0.26 ± 0.14 | 5.5E-04, 0.83 ± 0.47 | 2.6E-05, 1.21 ± 0.56 |
| 5q13.2<sup>†</sup> | 1.1E-03, -0.12 ± 0.07 | **rs609659 (G)**<br>**4.6E-08, -0.39 ± 0.14** | 1.6E-06, -0.47 ± 0.19 | 6.9E-04, -0.50 ± 0.29 | 1.2E-04, 0.32 ± 0.16 | 2.1E-03, 0.29 ± 0.18 | 4.1E-03, 0.28 ± 0.19 | 3.4E-03, -0.47 ± 0.32 |
| 7q22.1<sup>†</sup> | 4.9E-04, 0.16 ± 0.09 | 1.8E-03, 0.32 ± 0.2 | 5.2E-03, 1.33 ± 0.93 | 1.8E-04, 1.17 ± 0.61 | 1.7E-03, 0.20 ± 0.13 | 2.2E-03, 0.27 ± 0.17 | **rs6465810 (C)**<br>**1.2E-08, 1.17 ± 0.4** | rs6465810 (C)<br>5.7E-07, 0.78 ± 0.3 |
| 8q21.3<br>(*DCAF4L2*) | **rs12543318 (C)**<br>**5.4E-10, 0.22 ± 0.07** | 3.6E-05, 0.27 ± 0.13 | 7.6E-04, -0.33 ± 0.19 | 1.1E-03, 0.44 ± 0.26 | rs12543318 (C)<br>2.9E-08, 0.22 ± 0.08 | 4.5E-06, 0.21 ± 0.09 | 8.8E-06, 0.39 ± 0.17 | 6.2E-06, 0.32 ± 0.14 |
| 8q24.21(*p-ter*) | **rs7839784 (T)**<br>**2.2E-14, 0.34 ± 0.08** | rs55768865 (G)<br>5.2E-09, 0.88 ± 0.29 | rs55768865 (G)<br>5.1E-07, 1.04 ± 0.4 | 1.9E-05, 1.04 ± 0.47 | rs5894949 (A)<br>2.2E-11, 0.33 ± 0.09 | rs55768865 (G)<br>5.6E-10, 0.56 ± 0.17 | 2.8E-06, 0.54 ± 0.22 | rs55768865 (G)<br>1.9E-07, 0.84 ± 0.31 |
| 8q24.21(*q-ter*) | **rs72728755 (A)**<br>**3.1E-32, 0.58 ± 0.09** | rs72728755 (A)<br>6.4E-13, 0.72 ± 0.19 | rs112704402 (A)<br>1.3E-08, 0.74 ± 0.25 | rs72728755 (A)<br>1.5E-08, 0.91 ± 0.31 | rs72728755 (A)<br>1.5E-26, 0.59 ± 0.1 | rs72728755 (A)<br>8.6E-20, 0.58 ± 0.12 | rs72728755 (A)<br>1.7E-14, 1.04 ± 0.26 | rs72728755 (A)<br>1.2E-16, 0.89 ± 0.2 |
| 15q24.2-q24.1<br>(*ARID3B*) | **rs58691516 (CT)**<br>**6.7E-07, -0.20 ± 0.08** | 9.0E-06, -0.40 ± 0.17 | 1.2E-03, 0.35 ± 0.21 | 3.2E-04, -0.39 ± 0.21 | 4.0E-05, -0.19 ± 0.09 | 5.7E-06, -0.24 ± 0.1 | 1.2E-04, -0.48 ± 0.24 | 9.2E-04, -0.30 ± 0.18 |
| 17p13.1 (*NTN1*) | rs12944377 (C)<br>6.3E-09, -0.22 ± 0.07 | 1.5E-04, -0.29 ± 0.15 | 1.6E-04, -0.39 ± 0.2 | 3.3E-03, 0.32 ± 0.21 | rs12944377 (C)<br>1.7E-09, -0.26 ± 0.08 | **rs12944377 (C)**<br>**5.8E-11, -0.31 ± 0.09** | 5.8E-04, 0.75 ± 0.43 | 1.6E-04, 0.29 ± 0.15 |
| 17q21.31-q21.32<br>(*WNT9B:WNT3*) | **rs7216951 (T)**<br>**3.0E-07, -0.22 ± 0.08** | 9.2E-04, -0.40 ± 0.24 | 3.5E-03, -0.31 ± 0.21 | 5.4E-04, -0.60 ± 0.34 | rs7216951 (T)<br>4.8E-07, -0.25 ± 0.09 | 3.6E-06, -0.26 ± 0.11 | 7.3E-04, -0.52 ± 0.3 | 1.4E-03, 0.23 ± 0.14 |
| 17q23.3-q23.2<br>(*TANC2*) | 2.6E-06, -0.22 ± 0.09 | 1.2E-04, 0.30 ± 0.15 | **rs17683292 (C)**<br>**1.0E-07, 0.56 ± 0.2** | 1.3E-04, -0.42 ± 0.22 | 1.7E-05, -0.23 ± 0.1 | 6.4E-06, -0.23 ± 0.12 | 7.1E-04, -0.73 ± 0.42 | 1.8E-03, -0.58 ± 0.36 |
| 19p13.3<sup>†</sup> | 1.5E-03, 0.33 ± 0.2 | 6.4E-04, 0.25 ± 0.14 | 1.1E-03, 0.76 ± 0.46 | 2.9E-03, 0.39 ± 0.26 | 1.4E-04, 0.44 ± 0.23 | 2.4E-03, 0.41 ± 0.27 | **rs628271 (C)**<br>**1.9E-08, 1.68 ± 0.58** | 1.7E-04, 0.92 ± 0.48 |
| 20q13.33<sup>†</sup> | 2.2E-04, -0.30 ± 0.16 | 1.3E-04, 0.85 ± 0.43 | **rs2427238 (G)**<br>**1.5E-09, 1.94 ± 0.62** | 4.6E-05, 0.89 ± 0.42 | 4.4E-05, -0.37 ± 0.18 | 8.2E-05, -0.40 ± 0.2 | 5.0E-04, 0.83 ± 0.47 | 3.4E-04, 0.68 ± 0.37 |

Note: For each locus and GWAS, meta-analysis p-value, beta estimate and its 95% CI are shown; RS numbers and their effect alleles
(in parentheses) are shown for suggestive and significant associations - SNPs with the most significant p values at a locus may differ
across the GWASs,; <sup>†</sup>novel loci; p-values $\leq 5.0\text{E-}08$ highlighted in dark green and p-values ≤ $1.0\text{E-}06$ in light green; smallest p-value across subtypes highlighted in bold; two distinct association peaks in 8q24.21 locus listed separately.

The five novel associations observed are: (i) 3q29, most significantly associated with the CL_[CL+CLP]_ subtype, (ii) 5q13.2, most significantly associated with the CL_[CL+]_ subtype, (iii) 7q22.1 showing the strongest association with the CLP _[CL+CLP]_ subtype, (iv) 19p13.3 also showing the strongest association with the CLP_[CL+CLP]_ subtype, and (v) 20q13.3, associated with the CL_[CL]_ subtype.

The known OFC loci recapitulated here include the genes *PAX7, IRF6, FAM49A, DCAF4L2, ARID3B, NTN1, WNT9B:WNT3, TANC2*, and the 8q24.21 locus. Among these, *PAX7, FAM49A, DCAF4L2, ARID3B*, and *WNT9B:WNT3* are associated with both CL and CLP. The *IRF6* locus is the most strongly associated with the CL_[POFC]_ subtype, *TANC2* with the CL_[CL]_ subtype, and *NTN1* with CLP_[CLP]_ subtype. The 8q24.21 locus has traditionally been treated as a single locus, however, the prior CL/P GWAS study using samples from Pitt-OFC reported two distinct peak regions with genome-wide significant association p-values (Leslie et al. [12]). In the current study, we also observed two distinct peak regions at this locus. Both peaks are most strongly associated with the CL/P_[POFC]_ subtype.

### Identification of loci associated with specific cleft and/or family subtypes

Based on the strength of association and location of the most significant variants across subtypes, six previously reported OFC loci, *PAX7, FAM49A, DCAF4L2*, the 8q24.21 locus, *ARID3B, WNT9B:WNT3* and a novel locus 7q22.1 appear to be associated with both CL and CLP, i.e., the CL/P_[POFC]_ meta p-values were the most significant at these loci with subtypes represented by the larger samples - CLP_[CLP+]_ and CLP_[CLP]_ - produced more significant association p-values as compared to the subtypes with smaller samples. The remaining nine loci produced more significant p-values within a cleft or a family subtype. We hypothesized that the differences in p-values could be the result of the sample size differences between phenotypic subtypes. We therefore compared the estimated meta-analysis effect sizes of the associated variants within each of 15 peak regions identified above obtained for the eight phenotypes. This was done to verify whether the degree of risk for developing an OFC differed by OFC type and/or family type.

Table 2 lists the estimated beta coefficients and 95% confidence intervals for the top associated variant at each locus and for each subtype GWAS. The comparison showed statistically significant differences between the meta-analysis beta coefficients between subtypes at five of the associated loci, both between cleft subtypes (i.e. CL[_CL+]_ vs. CLP_[CLP+]_) and between family subtypes (i.e. CL_[CL]_, CLP_[CLP]_, CL_[CL+CLP]_ and CLP_[CL+CLP]_). A comparison of the ancestry-specific beta coefficients also showed variation similar to the meta-analysis effect sizes. A comparison of the frequency of the effect allele within affected individuals included in the phenotypic subsets showed that subtype-specific variants occurred at varying frequencies between subgroups. Overall, case allele frequencies were observed to differ between subtypes if effect sizes varied between subtypes, and vice versa.

Three of the loci considered as being associated with a specific subtype, are presented in figures 2-4 below. Fig 2 shows the *IRF6* locus; Fig 3 and Fig 4 show two interesting novel loci - 20q13.33 and 3q29; each containing multiple variants associated with genome-wide significant and/or suggestive p-values. These three figures illustrate that subtype-specific differences in strength of association mostly correspond to effect size differences, and also to differences in frequency of the effect allele amongst affected subjects (referred to as case EAFs) belonging to these subtypes. Differences in effect sizes and case EAFs that are observed at the meta-analysis level are also seen within ancestry groups, especially the two largest ones - CSA and EUR.

**Fig 2.**
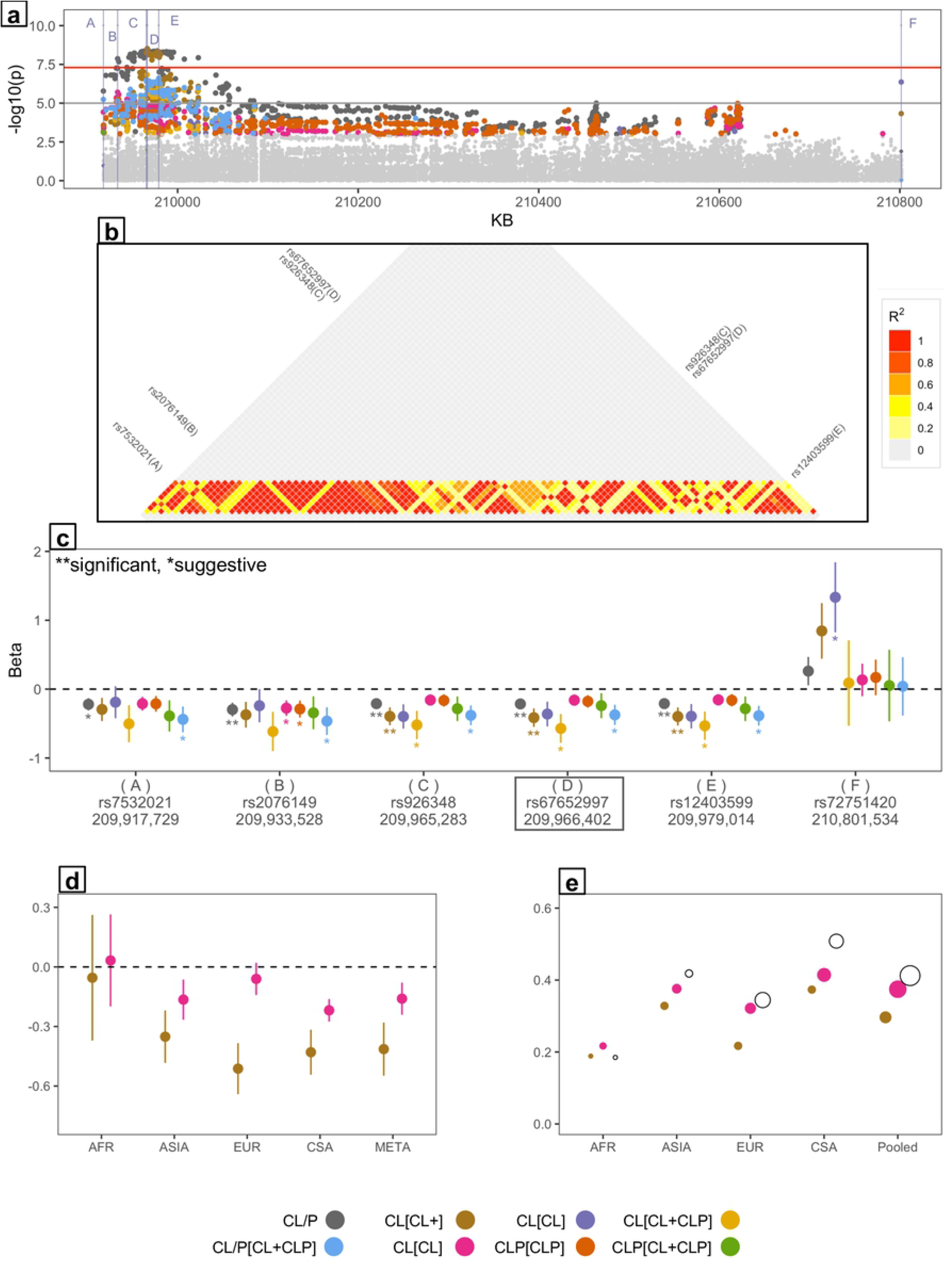
*IRF6* locus specific to CL_[CL+]_ subtype (a) regional Manhattan plot consisting of six distinct variants (A-F) with the most significant p-value from each subtype; (b) LD r^2^ values > 0.2 between variants (A-E) with p-value below 0.001, variant F is in a separate LD block from the A-E; (c) beta coefficient and 95% CI for variants A-F, D: lead variant at this locus, ** significant and * suggestive associations; (d) effect sizes and (e) effect allele frequency within affected subjects for cleft subtypes CL_[CL+]_ vs. CLP_[CLP+]_ by ancestry-subgroup.

**Fig 3.**
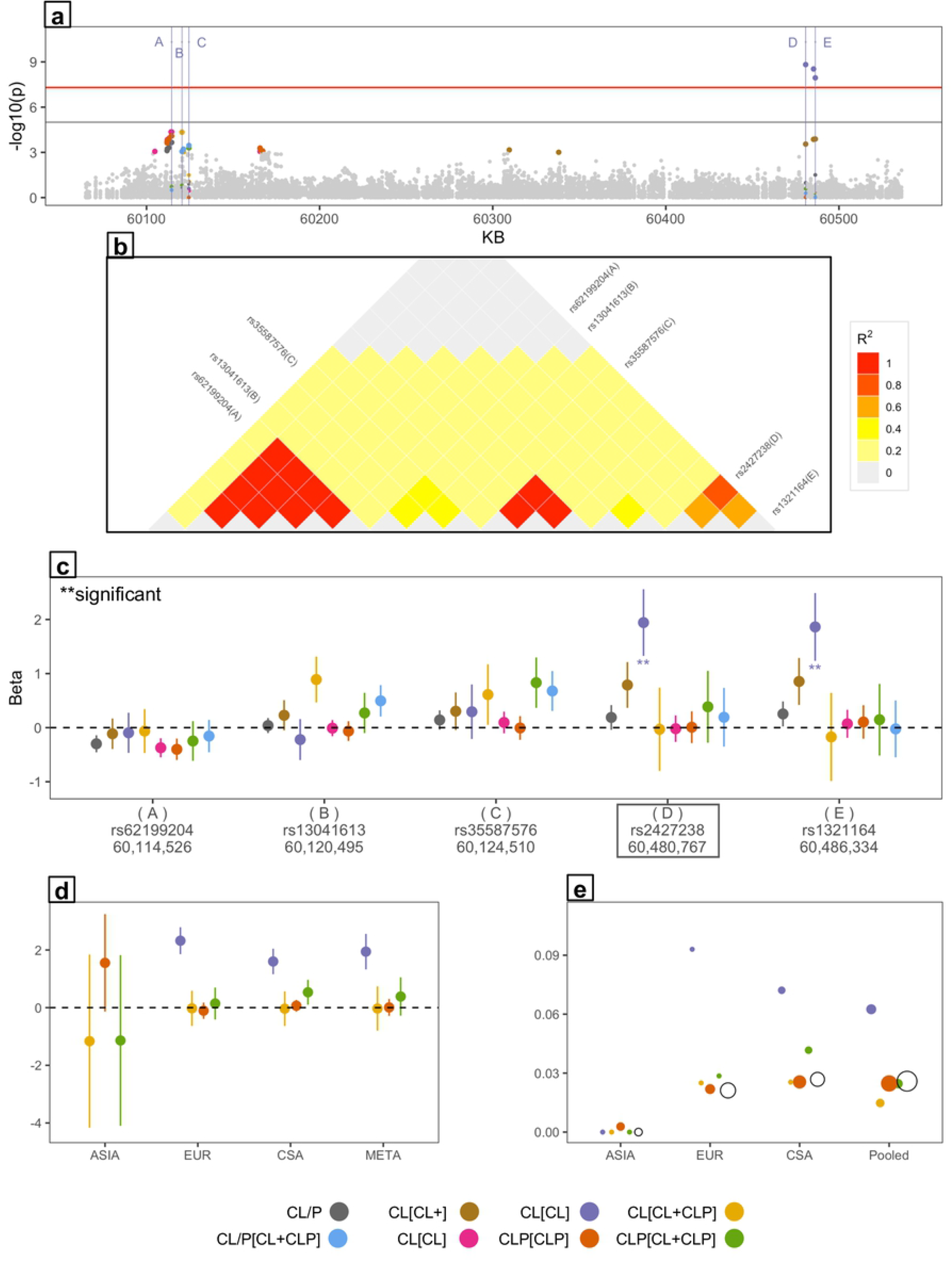
20q13.3 novel locus specific to CL_[CL]_ subtype (a) regional Manhattan plot consisting of five distinct variants (A-E) with the most significant p-value from each subtype; (b) LD r^2^ values > 0.2 between variants (A-E) with p-value below 0.001; (c) beta coefficient and 95% CI for variants A-E, D: lead variant at this locus, ** significant associations; (d) effect sizes and (e) effect allele frequency within affected subjects for family subtypes CL_[CL]_, CL_[CL+CLP]_, CLP_[CLP]_ and CLP_[CL+CLP]_ by ancestry-subgroup.

**Fig 4.**
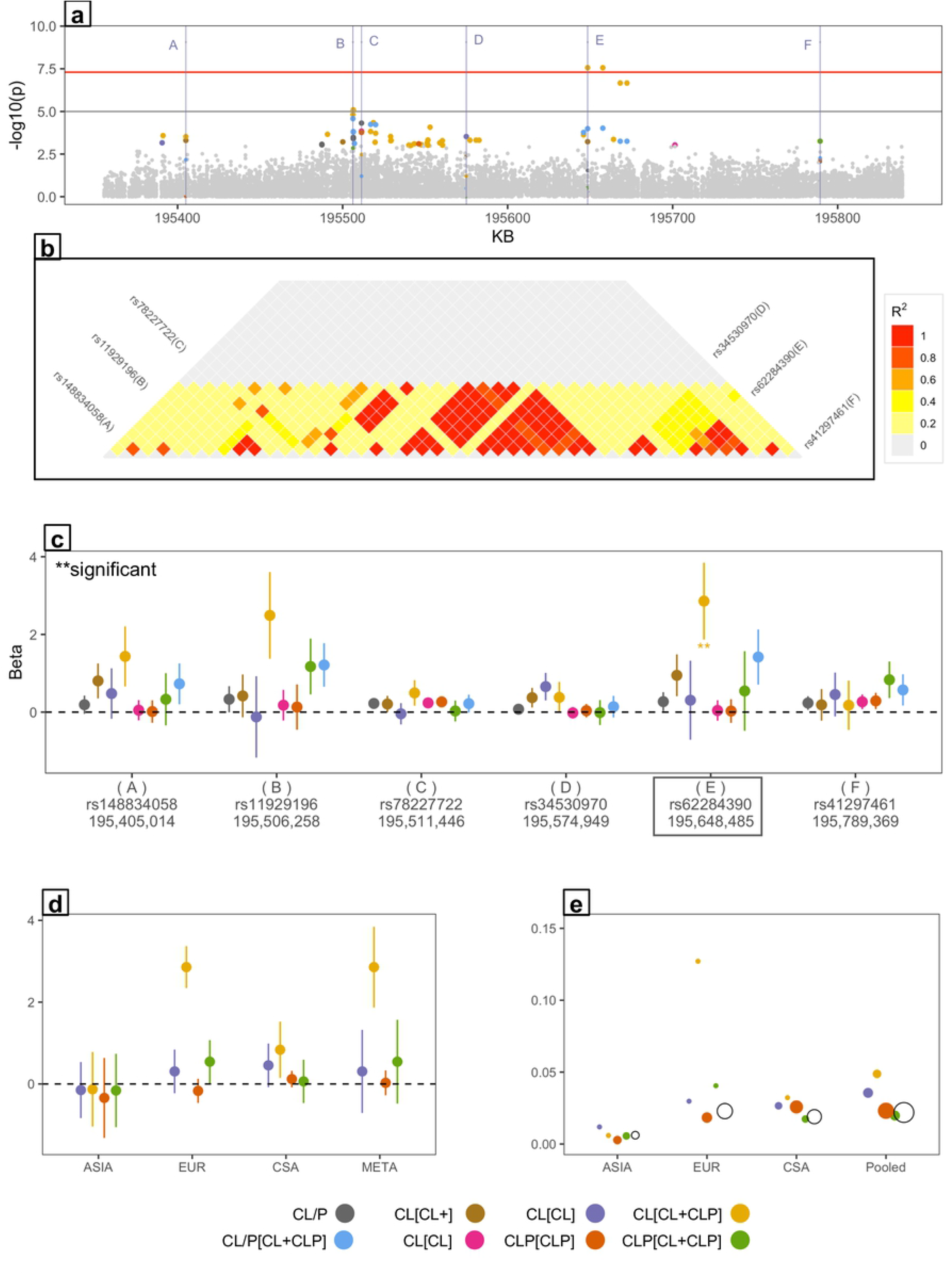
3q29 novel locus specific to CL_[CL+CLP]_ subtype (a) regional Manhattan plot consisting of six distinct variants (A-F) with the most significant p-value from each subtype; (b) LD r^2^ values > 0.2 between variants (A-F) with p-value below 0.001; (c) beta coefficient and 95% CI for variants A-F, E: lead variant at this locus, ** significant associations; (d) effect sizes, and (e) effect allele frequency within affected subjects for family subtypes CL_[CL]_, CL_[CL+CLP]_, CLP_[CLP]_ and CLP_[CL+CLP]_ by ancestry-subgroup.

In each figure, the top panel (a) shows a regional Manhattan plot with the most significant association per subtype – the top associations are labelled in order of their genomic position. Panel (b) in each figure shows the LD pattern of variants with p-value below 0.001 as that locus - LD r^2^ values above 0.2 shaded as indicated, and top associations labelled as in panel (a). Overall, LD patterns between top associations from the subtypes are as expected, i.e. LD is high between subtype-specific associations that are in close proximity, low (> 0.2) otherwise. Panel (c) shows the effect size estimates (beta coefficient and 95% CI) for the labelled associations for all subtypes – effect size estimates of significant and suggestive associations are identified in the forest plot, and the lead SNP name outlined. Panel (d) compares ancestry-subgroup specific effect sizes for either the two cleft subtypes (CL_[CL+]_ and CLP_[CLP+]_), or the four family subtypes (CL_[CL]_, CLP_[CLP]_, CL_[CL+CLP]_, CLP_[CL+CLP]_) at the lead SNP depending on which comparisons indicated subtype specificity. Panel (e) compares effect allele frequency within affected subjects in each subtype to that of controls at the lead SNP by ancestry. The observed variation in effect sizes across subtypes corresponds to differences in case EAFs, i.e. case EAFs within subtypes differ from one another, if the effect sizes are different, with a single exception – the 5q13.2 locus, which is further explored in the next section.

#### 1. Loci specific to the CL cleft-subtype

The novel locus at **5q13.2**, and the known **1q32.2 (*IRF6*)** locus show the most significant association for the CL_[POFC]_ cleft subtype. Fig 2 shows the *IRF6* locus in detail: the regional Manhattan plot (Fig 1a) shows six distinct variants (labelled A-F) with the most significant p-values from the subtype meta-analyses. The top association for CL_[CL+]_ coincides with the top CL_[CL+CLP]_ variant (SNP D: rs67652997 in Fig 2c), although the latter shows lower significance, and the top associations for CLP_[CLP+]_ and CLP_[CLP]_ also coincide (SNP B: rs2076149). LD between variants with significance p-values (below 0.001) is shown for the 209.92-209.98 KB region spanning five of these variants (A-E); the top CL_[CL]_ association is not shown - it is in low LD with the rest of the top associations.

The largest CL effect size is observed for the CL_[CL+]_ subtype, as can be seen in Fig 2c for *IRF6*. The CL_[CL+]_ subtype’s effect sizes at the lead SNP rs609659, as well as nearby variants in LD with the lead SNP is distinctly larger in magnitude than for the CLP_[CLP+]_ subtype. Effect sizes for the CL_[CL]_ and CL_[CL+CLP]_ family-based subtypes are also larger than the CLP_[CLP]_ and CLP_[CL+CLP]_ effect sizes, while CL_[CL]_ and CL_[CL+CLP]_ effect sizes are not statistically different. These loci show stronger association to CL, attributable to both the CL_[CL]_ and CL_[CL+CLP]_ family subtypes. Within the *IRF6* gene, the lead variant is observed to have a protective effect on CL risk and observed at a lower frequency than the non-effect allele within cases in EUR and CSA. Within ASIA and AFR, effect sizes appear to be similar between CL_[CL+]_ and CLP_[CLP+]_. At the 5q13.2 locus, the ancestry subgroup-specific effect sizes are consistent with the meta-analysis effect sizes within the ASIA, EUR and CSA subgroups, i.e. CL_[CL+]_ effect sizes are larger in magnitude than CLP_[CLP+]_. Beta coefficients overlap within the AFR subgroup. The EAF within CL_[CL+]_ affecteds of all ancestries pooled is not different from the EAF in CLP_[CLP+]_ cases, unlike variants within the other subtype-specific loci. However, this appears to be due to EAF differences across ancestry groups: in AFR, the CL_[CL+]_ EAF is smaller than the CLP_[CLP+]_, while the reverse is true in ASIA, EUR and CSA (supplement Fig S1).

#### 2. Loci specific to the CL_[CL]_ family-subtype

At two peak regions, the novel locus at 20q13.33, and 17q23.2;q23.3 (*TANC2*), the CL_[CL]_ meta-analysis p-value is the most significant, and the CL_[CL]_ meta-analysis effect sizes are much larger than the other family-type based subsets. Notably, the CL_[CL+]_ effect size is not different from the CLP_[CLP+]_ subtype. Fig 3 highlights the main association outcomes at the 20q13.33 locus. As seen in Fig 3d, the variation in beta estimates within the CSA and EUR subgroups correspond to the variation observed within the overall meta-analysis beta estimates, and the lead variant for CL_[CL]_ shows a positive effect size (beta), while other effect sizes are close to zero. The effect allele was not observed in CL_[CL]_ families from ASIA, and AFR was excluded from the family-subtype comparison (Fig 3e). At the other locus showing association within the CL_[CL]_ subtype - *TANC2*, effect size differences were observed in the EUR and CSA group, with differences observed in the ASIA group. Further, within the CSA group, the CL_[CL]_ subtype showed a positive effect whereas the CL_[CL+CLP]_ subtype showed a negative effect, which was not the case for EUR. EAFs within the affecteds were consistently highest in the CL_[CL]_ subtype sample than the other family-subtypes, and the effect allele is least frequent in ASIA (Supplement Fig S2).

#### 3. 3q29 locus specific to CL_[CL+CLP]_ family-subtype

The **3q29 novel locus** is more strongly associated with the CL_[CL+CLP]_ subtype than any other subtype (Fig 4). There is low LD between SNPs associated with different subtypes as seen in Fig 4b. The CL_[CL+CLP]_ subtype’s effect size is much larger than that of other subtypes also resulting in a significant difference between the CL_[CL+]_ subtype’s effect size and the CLP_[CLP+]_ subset’s effect size (Fig 4c and 4d). The **3q29** locus is another instance where ancestry plays a role. The elevated beta in CL_[CL+CLP]_ is due to samples of EUR ancestry, and the corresponding EAF in the EUR subgroup is also much higher than EAFs of other family subtypes (Fig 4e). Effect size variation is not observed in CSA, which is consistent with similar case EAFs in CSA, and the effect allele is very rarely observed in ASIA. When effect sizes from the ancestry-based subgroups are examined, the difference between CL_[CL]_ and CL_[CL+CLP]_ effect sizes is observed in the EUR subgroup, but not in ASIA and CSA.

#### 4. Locus specific to CLP_[CL+CLP]_ family-subtype

The **19p13.3 peak** includes a single Bonferroni-significant association at SNP rs628271; with no other neighboring variants reaching a suggestive level of significance, this may not be a reliable association. Even so, interestingly the effect size of this variant for the CLP_[CL+CLP]_ subtype is larger than all the other family-based subtypes. The CL_[CL+]_ subtype effect size is similar to the CLP_[CLP+]_ effect size. This difference is observed in CSA and EUR, but not in ASIA.

#### 5. Loci with no variation in subtype-specific effect sizes

At the following loci, the subtype-specific effect sizes are similar in magnitude and direction to those from the other subtypes, indicating that that these loci affect the risk of both CL and CLP to a similar extent regardless of family classification: 1p36.13 (*PAX7*), 2p24.2-24.3 (*FAM49A*), 7q22.1 - novel locus, 8q21.3 (*DC4FL2*), both peaks within 8q24.1, 15q24.1;q24.2 (*ARID3B*), 17p13.1 (*NTN1*), and 17q21.31;q21.32 (*WNT9B;WNT3*). At these loci, larger samples yielded more significant association p-values.

## DISCUSSION

For the five novel loci observed in our study, a bioinformatics search yielded interesting, but not conclusive indication of their roles in the development of OFCs. The lead variant within 5q13.2 is in close proximity to the *TMEM1* gene, and the lead variant within the 20q13.33 locus is intronic to the *CDH4* gene; both *TMEM1* and *CDH4* are involved in the Wnt signaling pathway, known to be involved in the development of OFCs. The lead variant in our 3q29 locus is located approximately 1 MB downstream of the *DLG1* gene, reported as being associated with CL/P in a recent study of CL/P on a Polish population [23]. In our study, however, we observed only weak association to variants within the *DLG1* gene. The other three loci contain craniofacial super-enhancer regions. The top associations in the 7q22.1 locus are intronic to the *COL26A1* and *RANBP3* genes, both reported as having a blood phenotype (UCSC genome browser, https://genome.ucsc.edu/index.html). It is interesting to note that the previously reported genome-wide linkage and targeted region study of Pitt-OFC pedigree subsets based on cleft types [21] reported two regions – 9q21.33 and 14q21.3 – that were associated at a suggestive level of significance in our study, although the current associations do not lie within the fine-mapped regions analyzed in the former study.

The analysis of CL and CLP as a single phenotype (CL/P) in the [CL+CLP] families did not produce unique associations, as would be expected if these families were segregating for genes that cause a continuum of the CL/P phenotype. This lack of association may further support the hypothesis that CL/P is not a single phenotype etiologically. Further, we hypothesize that our family subtype-based analyses show evidence of genetic heterogeneity even within the cleft subtypes CL and CLP themselves. For example, association of CL to *TANC2* is much stronger in the [CL] families than in the [CL+CLP] families, while the reverse is true at the 3q29 locus. Finally, our study outcomes show consistently stronger and more reliable associations for the CL-based subtypes (5 previously known and novel loci) as compared to the CLP-based subtypes (a single novel locus), although the sample sizes for the CLP-based subtypes are larger. Our study results recapitulated the association of *IRF6* with CL [24]. We thus hypothesize that CL is genetically more homogeneous than CLP. A possible alternative to genetic heterogeneity would be phenotypic heterogeneity: there exists diagnostic uncertainty with the palate phenotype, it is sometimes left undiagnosed, or, in some cases, the presence of submucous CP along with CL is not categorized as CLP. However, Pitt-OFC subjects were thoroughly examined for submucous CP and VPI, so this would be unlikely to have happened on large enough scale to impact our analysis outcomes.

This study makes an important contribution to the study of heterogeneity between OFC types using a study design where both the individuals as well as the family’s OFC types are incorporated. The idea that genetically related individuals also tend to have the same type of OFC more often than different types of OFCs (REF), has been rarely utilized in running GWASs of OFC subtypes. Our study provides a methodology for incorporating the proband’s relatives’ cleft types within the GWAS framework, and the observed outcomes provide valuable insight into etiological differences between OFC subtypes.

## METHODS

### Study sample

Our study sample consists of participants from the multiethnic Pittsburgh Orofacial Cleft study (Pitt-OFC) [12], including a variety of pedigree structures and sizes, and including both simplex as well as multiplex families. Sample recruitment was carried out in accordance with ethics approval procedures at the University of Pittsburgh, the coordinating center for the Pitt-OFC study, as well as the respective institutions that contributed samples to the Pitt-OFC study. Genotyping was carried out at the Center for Inherited Disease Research (CIDR) at Johns Hopkins University, on an Illumina chip for approximately 580,000 variants genome-wide as summarized previously [12, 13], and available from dbGaP (**dbGaP Study Accession:** phs000774.v2.p1). The CIDR coordinating center at the University of Washington was also responsible for ensuring the quality of called genotypes. Subsequently, genotypes were imputed using the “1000 genome project phase 3” reference panel, at approximately 35,000,000 variants of the GrCH37 genome assembly. Genotyping, quality control, and imputation steps were previously described in detail in Leslie et al. [12].

The full sample – POFC – utilized in our current study includes 2,218 individuals affected with CL or CLP, and 4,537 unaffected relatives from 1,939 families that contain members affected with CL and/or CLP. The types of OFCs present in a pedigree were obtained by direct participation by affected individuals and/or by a reported family history of OFCs. An additional 2,673 unaffected individuals from 1,474 families with no reported history of an OFC (referred to as Controls) are included in the association analysis. Participants from pedigrees containing individuals affected with a cleft palate only (CP), or having a reported family history of CP were excluded from this study.

### Definition of subtypes

Several subsets were created from the POFC sample based on the types of OFCs reported within pedigrees, as follows. First, the pedigrees were partitioned into three non-overlapping subsets, (i) [CL]: pedigrees that contain individuals affected with CL only, but not members affected with CLP, (ii) [CLP]: pedigrees that contain individuals affected with CLP but not members affected with CL only, and (iii) [CL+CLP]: pedigrees containing some members affected with CL only as well as some members affected with CLP. The partitioning of pedigrees into these three subsets used all available phenotypic and relationship information, including phenotypic information from pedigree members who were not genotyped. Two additional subsets were then defined, (iv) [CL+], all pedigrees with any CL-affected member, i.e. the union of [CL] and [CL+CLP], and (v) [CLP+], all pedigrees with any CLP-affected member, i.e. the union of [CLP] and [CL+CLP]. The [CL+] and [CLP+] subsets are not disjoint, i.e. they both contain subjects from [CL+CLP] pedigrees.

Eight GWAS phenotypic subtypes were then defined for these five subsets of pedigrees for running genome-wide association analysis, and affection statuses assigned to pedigree members belonging to each of the eight phenotypic subtypes as described below. The 2,673 Controls were included in each of the GWASs.

A. **CL/P**_**[POFC]**_ – Within the full POFC sample, participants with either a CL, or CLP were set to affected, participants without any OFC were set to unaffected.
B. **CL**_**[CL]**_ –Within the [CL] pedigrees - group (i) above, participants with CL were set to affected, and those without CL were set to unaffected.
C. **CLP**_**[CLP]**_ – Within the [CLP] group of pedigrees – group (ii), participants with CLP were set to affected, and those without CLP were set to unaffected.
D. **CL/P**_**[CL+CLP]**_ – within the [CL+CLP] group of pedigrees – group (iii), participants with either CL or CLP were set to affected, and those without OFCs were set to unaffected.
E. **CL**_**[CL+CLP]**_ –Within [CL+CLP] pedigrees – group (iii), participants with a CL only were set to affected, those with CLP were set to unknown (thereby excluding them from GWAS), and those without OFCs were set to unaffected.
F. **CLP**_**[CL+CLP]**_ –Within [CL+CLP] pedigrees – group (iii), pedigree members with a CLP were set to affected, those with CL only were set to unknown (thereby excluding them from GWAS), and those without OFCs were set to unaffected.
G. **CL**_**[CL+]**_ – Within the [CL+] – group (iv) pedigrees, participants with CL only were set to affected, those with CLP were set an unknown affection status (thereby excluding them from GWAS), and those without any OFC were set to unaffected.
H. **CLP**_**[CLP+]**_ – Within the [CLP+] pedigrees – group (v), participants affected with CLP were set to affected, those with CL only were set to unknown (thereby excluding them from GWAS), and those without any OFC were set to unaffected.

Fig 1 shows the partitioning of POFC pedigrees into the eight phenotypic subsets and phenotype definitions within each of these phenotypic subsets that were used to run separate GWASs. For illustration purposes, each subtype is depicted as simple nuclear pedigree structures with three offspring, two of which are affected with CL or CLP, although a wide variety of family types are represented in this study. Simplex and multi-generation pedigrees were handled following the same procedure for grouping into subtypes. In addition to the type of pedigrees included in each subset, Fig 1 also depicts affected and unaffected members, as well as those assigned an unknown affection status, thereby excluding these members from the corresponding GWAS.

### Genome wide association

We have shown previously that the degree of OFC risk at certain susceptibility loci varies with ancestry of the sample participants [22]. In order to control for this variance, we first classified subjects into four different genetically defined ancestry groups using the principal component analysis-based classification defined in a previous study using POFC subjects [12]. For each of the eight GWAS phenotypic samples defined above and shown in Fig 1, we first analyzed each ancestry group separately, then combined the association outcomes using meta-analysis. The four ancestry-based groups were: AFR (participants of African origin), ASIA (participants of Asian origin), EUR (those of European white origin), and CSA (participants of Central and Southern American origin). Table 3 shows the breakdown of the analysis sample by ancestry, pedigree type, and affection status.

**Table 3.**
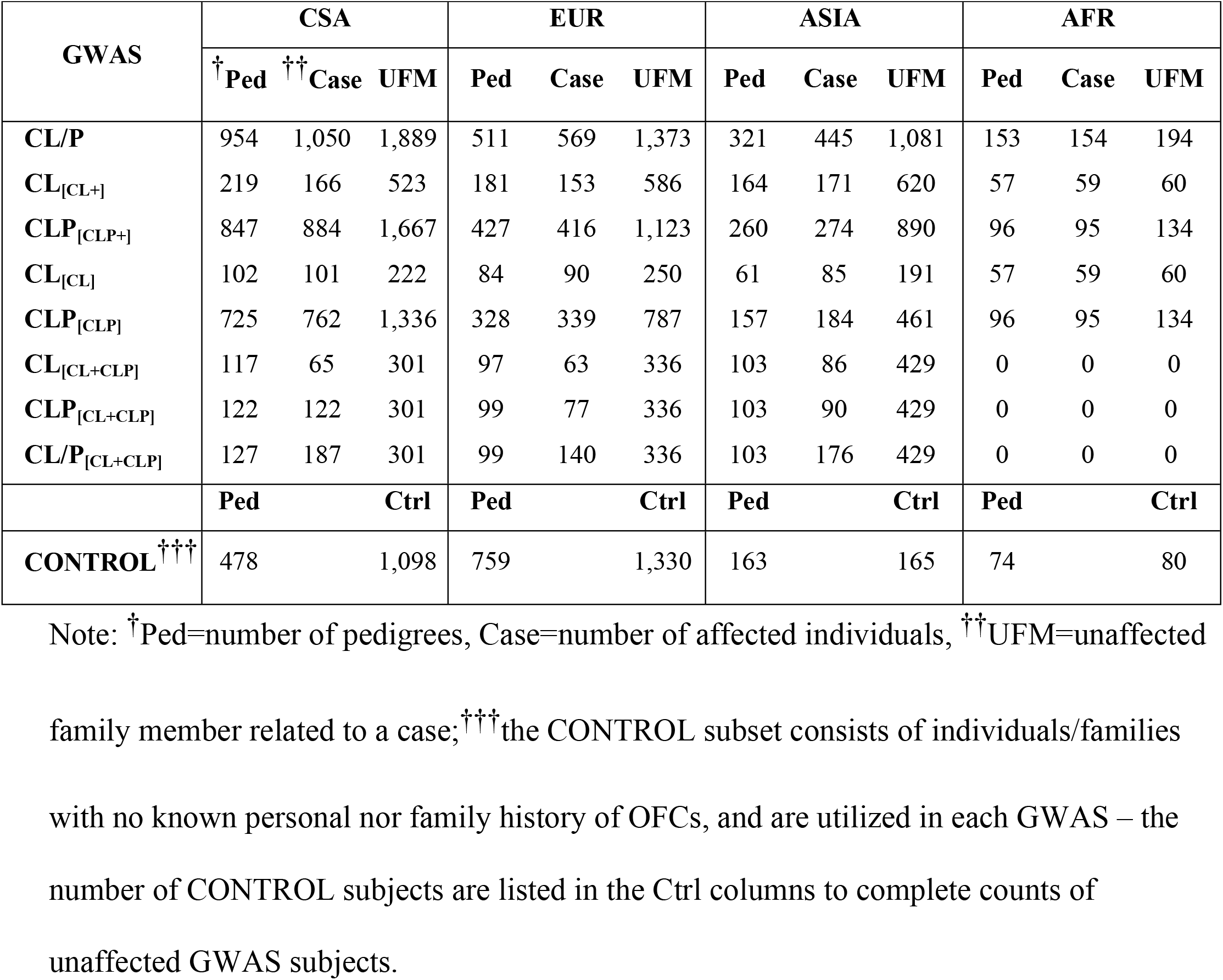
Counts of pedigrees and participants by GWAS, ancestry and affection status

Individual GWASs were run using the mixed-model association program, GENESIS [25]. GENESIS uses a genetic relationship matrix (GRM) estimated from the observed genotype data to account for population structure and familial relatedness, therefore, it is not necessary to correct for population admixture using ancestry PCs. The use of a GRM is necessary to account for population admixture within our ancestry-based subsets, which, in turn is due to the varying geographical origin of participants in each of these subsets (see Supplementary Table S2 for a breakdown by recruitment site). The genetic relationship matrix also provides an estimate of the polygenic variance component. Significance of association is based on the score test, comparing the maximum likelihood of disease outcomes conditional on observed genotypes at each variant to the maximum likelihood of the unconditional polygenic model. GENESIS reports approximate effect sizes in the form of betas, i.e. the log-likelihood ratio of the conditional and unconditional model) and standard error of the effect size. In this study, the effect allele is fixed across all GWASs as the minor allele at each variant identified in the combined POFC sample.

Ancestry-specific GWASs were then meta-analyzed for each of the eight GWAS phenotypes using the inverse-variance method implemented in PLINK [26]. The reported odds ratios from PLINK were converted to log-scale effect sizes, to conform to the GENESIS reported effects. The 95% confidence intervals of betas were calculated under the assumption that the meta-analysis p-values are distributed normally. All four ancestry-groups were meta-analyzed for the CL_[CL+]_ and CLP_[CLP+]_ subtypes. There are no AFR pedigrees containing both CL and CLP affected members, therefore, meta-analysis was conducted excluding the African samples (AFR) for the five family-subtypes (CL_[CL]_, CL_[CL+CLP]_, CLP_[CLP]_, CLP_[CL+CLP]_ and CL/P_[CL+CLP]_).

### Variant selection

Genotyped and imputed variants that passed quality control, and had minor allele frequencies of 2% or more within their respective GWAS sample subsets were used to run association. The observed minor allele frequencies of reported loci were checked against values obtained from the gnomAD database [27] to guard against imputation inaccuracy.

### Identification of novel associations

For each genome-wide meta-analysis, variants showing association p-values below 1.0E-06 were selected for further investigation, and grouped into association peaks measuring 1MB or less. We then checked for overlap between our associations peaks with the 29 genomic regions listed as harboring known OFC genes by Beaty et al. [28] as well as associated regions reported by six recently published OFC GWAS studies. The six recent GWASs include (1) combined meta-analysis of parent-offspring trio and case-control cohorts from the current Pitt-OFC multiethnic study sample [12], (2) meta-analysis of the cohorts used in (1) with another OFC sample consisting of European and Asian participants [13], (3) GWAS of cleft lip with cleft palate in Han Chinese samples [18], (4) GWAS of cleft lip only and cleft palate only in Han Chinese [19], (5) GWAS of cleft lip with or without cleft palate in Dutch and Belgian participants [29] and (6) GWAS of sub-Saharan African participants from Nigeria, Ghana, Ethiopia and the Republic of Congo [30].

For each OFC gene, we checked if any our 1 MB association peaks overlapped with the span of the gene, as determined by its start and end transcription sites. The base pair positions for start and end transcription sites were obtained from the UCSC genome browser (https://genome.ucsc.edu/index.html) mapped to the February 2009 (GRCh37) assembly. For the 8q24.21 locus, which is a gene desert, we checked whether any of our associated SNPs were located in the 8q24.21 chromosome band. The distance between variants published by the six recent GWASs and our variants with p-values below 1.0E-06 were similarly measured, and a positive overlap reported if this distance was less than 500 Kb.

### Comparison of association outcomes between subtypes

Within each peak region the variant with the smallest meta-analysis association p-value observed for each of the eight subtypes were selected and their effect sizes compared. Effect size of each variant is represented by the beta coefficient of the SNP main effect under an additive model of inheritance, setting the minor allele (based on the entire POFC study sample) as the effect allele. Effect size and magnitude were compared across subtypes for the variants selected for each subtype to determine whether the 95% confidence intervals of effect size estimates overlapped. Next, LD r^2^ between selected variants at each locus was calculated using the PLINK program and the set of genotyped founders in the full POFC sample, irrespective of their OFC status. Finally, the observed effect allele frequency (EAF) within cases from the two GWASs were examined to assess whether these differed significant between cleft subtypes. We have previously shown that ancestry impacts association to CL/P in our POFC sample [22]; therefore, we examined the subtype-specific effect sizes within each ancestry group to assess whether the differences observed were similar to the those observed for the meta-analysis. EAFs within cases were also compared across the eight phenotypic subtypes within each ancestry group in addition to the cases pooled across ancestry groups for each phenotypic subset. In our study, we did not carry out a statistical test (e.g. Cochran’s Q statistic) to compare association outcomes from the OFC subtypes, as the unaffected relatives of OFC subjects and subjects from control families were used in the GWAS of more than one subtype; therefore, we relied mainly on qualitative evaluation of differences in the association outcomes.

## Data Availability

All data are fully available without restriction from dbGaP (dbGaP Study Accession: phs000774.v2.p1).

https://www.ncbi.nlm.nih.gov/projects/gap/cgi-bin/study.cgi?study_id=phs000774.v2.p1

## ACKNOWLEDGEMENTS

The authors wish to thank the participant families worldwide, without whom this research would not have been possible. Special thanks to Dr. Eduardo Castilla (deceases), Dr. Juan C. Mereb, Dr. Andrew Czeizel, and to the devoted staff at the many recruitment sites. This work was supported by grants from the National Institutes of Health including: X01-HG007485 [MLM], R01-DE016148 [MLM, SMW], U01-DE024425 [MLM], R37-DE008559 [JCM, MLM], R01-DE009886 [MLM], R21-DE016930 [MLM], R01-DE014667 [LMM], R21-DE016930 [MLM], R01-DE012472 [MLM], R01-DE011931 [JTH], U01-DD000295 [GLW], R00-DE025060 [EJL], R01- DE028342 [EJL], R01- DE28300 [AB]. Genotyping and data cleaning were provided via an NIH contract to the Johns Hopkins Center for Inherited Disease Research: HHSN268201200008I. Additional support provided by: an intramural grant from the Research Institute of the Children’s Hospital of Colorado [FWD]; operating costs support in the Philippines was provided by the Institute of Human Genetics, National Institutes of Health, University of the Philippines, Manila [CP]; grants through FAPERJ [IMO].

## Notes

### Competing Interest Statement

The authors have declared no competing interest.

### Author Declarations

University of Pittsburgh Internal Review Board

## BIBLIOGRAPHY

1. Nidey N, Moreno Uribe LM, Marazita MM, Wehby GL. Psychosocial well-being of parents of children with oral clefts. Child: care, health and development. 2016;42(1):42–50.

2. Wehby GL, Cassell CH. The impact of orofacial clefts on quality of life and healthcare use and costs. Oral diseases. 2010;16(1):3–10.

3. Berk NW, Marazita ML. The Costs of Cleft Lip and Palate: Personal and Societal Implications. In: Wyszynski DF, editor. Cleft Lip and Palate: From Origin to Treatment. Oxford: Oxford University Press; 2002.

4. Nidey N, Wehby G. Barriers to Health Care for Children with Orofacial Clefts: A Systematic Literature Review and Recommendations for Research Priorities. Oral Health and Dental Studies. 2019;2(1):2.

5. Naros A, Brocks A, Kluba S, Reinert S, Krimmel M. Health-related quality of life in cleft lip and/or palate patients - A cross-sectional study from preschool age until adolescence. Journal of cranio-maxillo-facial surgery:official publication of the European Association for Cranio-Maxillo-Facial Surgery. 2018;46(10):1758–63.

6. Bille C, Winther JF, Bautz A, Murray JC, Olsen J, Christensen K. Cancer risk in persons with oral cleft--a population-based study of 8,093 cases. American journal of epidemiology. 2005;161(11):1047–55.

7. Bui AH, Ayub A, Ahmed MK, Taioli E, Taub PJ. Association Between Cleft Lip and/or Cleft Palate and Family History of Cancer: A Case-Control Study. Annals of plastic surgery. 2018;80(4 Suppl 4):S178–s81.

8. Taioli E, Ragin C, Robertson L, Linkov F, Thurman NE, Vieira AR. Cleft lip and palate in family members of cancer survivors. Cancer investigation. 2010;28(9):958–62.

9. Dixon MJ, Marazita ML, Beaty TH, Murray JC. Cleft lip and palate: understanding genetic and environmental influences. Nature reviews Genetics. 2011;12(3):167–78.

10. Marazita ML, Leslie EJ. Genetics of Nonsyndromic Clefting. In: Losee J, Kirschner R, editors. Comprehensive Cleft Care. Second ed. Boca Raton, FL: CRC Press; 2016. p. 207–24.

11. Moreno Uribe LM, Marazita ML. Epidemiology, Etiology and Genetics of Orofacial Clefting. In: Shetye P, Gibson TL, editors. Cleft and Craniofacial Orthodontics: Wiley; In press.

12. Leslie EJ, Carlson JC, Shaffer JR, Feingold E, Wehby G, Laurie CA, et al. A multi-ethnic genome-wide association study identifies novel loci for non-syndromic cleft lip with or without cleft palate on 2p24.2, 17q23 and 19q13. Human molecular genetics. 2016;25(13):2862–72.

13. Leslie EJ, Carlson JC, Shaffer JR, Butali A, Buxo CJ, Castilla EE, et al. Genome-wide meta-analyses of nonsyndromic orofacial clefts identify novel associations between FOXE1 and all orofacial clefts, and TP63 and cleft lip with or without cleft palate. Hum Genet. 2017;136(3):275–86.

14. Harville EW, Wilcox AJ, Lie RT, Vindenes H, Abyholm F. Cleft lip and palate versus cleft lip only: are they distinct defects? American journal of epidemiology. 2005;162(5):448–53.

15. Grosen D, Chevrier C, Skytthe A, Bille C, Mølsted K, Sivertsen A, et al. A cohort study of recurrence patterns among more than 54,000 relatives of oral cleft cases in Denmark: support for the multifactorial threshold model of inheritance. J Med Genet. 2010;47(3):162–8.

16. Carlson JC, Anand D, Butali A, Buxo CJ, Christensen K, Deleyiannis F, et al. A systematic genetic analysis and visualization of phenotypic heterogeneity among orofacial cleft GWAS signals. Genetic epidemiology. 2019;43(6):704–16.

17. Carlson JC, Taub MA, Feingold E, Beaty TH, Murray JC, Marazita ML, et al. Identifying Genetic Sources of Phenotypic Heterogeneity in Orofacial Clefts by Targeted Sequencing. Birth defects research. 2017;109(13):1030–8.

18. Yu Y, Zuo X, He M, Gao J, Fu Y, Qin C, et al. Genome-wide analyses of non-syndromic cleft lip with palate identify 14 novel loci and genetic heterogeneity. Nature communications. 2017;8:14364.

19. Huang L, Jia Z, Shi Y, Du Q, Shi J, Wang Z, et al. Genetic factors define CPO and CLO subtypes of nonsyndromicorofacial cleft. PLoS genetics. 2019;15(10):e1008357.

20. Moreno Uribe LM, Fomina T, Munger RG, Romitti PA, Jenkins MM, Gjessing HK, et al. A Population-Based Study of Effects of Genetic Loci on Orofacial Clefts. Journal of dental research. 2017;96(11):1322–9.

21. Marazita ML, Lidral AC, Murray JC, Field LL, Maher BS, Goldstein McHenry T, et al. Genome scan, fine-mapping, and candidate gene analysis of non-syndromic cleft lip with or without cleft palate reveals phenotype-specific differences in linkage and association results. Hum Hered. 2009;68(3):151–70.

22. Mukhopadhyay N, Feingold E, Moreno-Uribe L, Wehby G, Valencia-Ramirez LC, Muneton CPR, et al. Genome-Wide Association Study of Non-syndromic Orofacial Clefts in a Multiethnic Sample of Families and Controls Identifies Novel Regions. Front Cell Dev Biol. 2021;9:621482.

23. Mostowska A, Gaczkowska A, Żukowski K, Ludwig KU, Hozyasz KK, Wójcicki P, et al. Common variants in DLG1 locus are associated with non-syndromic cleft lip with or without cleft palate. Clinical genetics. 2018;93(4):784–93.

24. Rahimov F, Marazita ML, Visel A, Cooper ME, Hitchler MJ, Rubini M, et al. Disruption of an AP-2alpha binding site in an IRF6 enhancer is associated with cleft lip. Nature genetics. 2008;40(11):1341–7.

25. Gogarten S, Sofer T, Chen H, Yu C, Brody J, Thornton T, et al. Genetic association testing using the GENESIS R/Bioconductor package. Bioinformatics. 2019.

26. Chang CC, Chow CC, Tellier LC, Vattikuti S, Purcell SM, Lee JJ. Second-generation PLINK: rising to the challenge of larger and richer datasets. GigaScience. 2015;4(1).

27. Karczewski KJ, Francioli LC, Tiao G, Cummings BB, Alföldi J, Wang Q, et al. Variation across 141,456 human exomes and genomes reveals the spectrum of loss-of-function intolerance across human protein-coding genes. 2019:531210.

28. Beaty TH, Marazita ML, Leslie EJ. Genetic factors influencing risk to orofacial clefts: today‘s challenges and tomorrow’s opportunities. F1000Research. 2016;5:2800.

29. van Rooij IA, Ludwig KU, Welzenbach J, Ishorst N, Thonissen M, Galesloot TE, et al. Non-Syndromic Cleft Lip with or without Cleft Palate: Genome-Wide Association Study in Europeans Identifies a Suggestive Risk Locus at 16p12.1 and Supports SH3PXD2A as a Clefting Susceptibility Gene. Genes. 2019;10(12).

30. Butali A, Mossey PA, Adeyemo WL, Eshete MA, Gowans LJJ, Busch TD, et al. Genomic analyses in African populations identify novel risk loci for cleft palate. Human molecular genetics. 2019;28(6):1038–51.

